# 365 days with COVID-19 in Iran: data analysis and epidemic curves

**DOI:** 10.1101/2021.03.02.21252694

**Authors:** Ebrahim Sahafizadeh, Saeed Talatian Azad

## Abstract

**Background:** The first confirmed cases of COVID-19 in Iran were reported on February 19, 2020. This study aimed to analyze the epidemic curves and to investigate the correlation between epidemic parameters and furthermore to analyze the impact of control measures on the spread of COVID-19 in Iran during 365 days of the epidemic.

**Methods:** We used data from February 20, 2020, to February 18, 2021, on the number of COVID-19 cases reported by Iranian governments. Pearson correlation coefficient was applied to investigate the correlation between different epidemic parameters. The number of daily deaths per daily new cases was averaged to calculate daily death rate and the same method was used to investigate the rate of daily positive tests. Furthermore, we employed two different methods to calculate the effective reproduction number using reported data.

**Results:** The results showed that there was a strong correlation between the number of daily deaths and the number of daily new cases (specially the admitted cases). The results also indicated that the mean of daily death rate of COVID-19 during 365 days was 4.9 percent, and averagely 13.9 percent of daily tests results were positive. Furthermore, epidemic curves showed that implementing strict social distancing measures effectively reduced the number of confirmed cases. The effective reproduction curve indicated that social distancing is still necessary to control the spread of COVID-19 in Iran.

**Conclusions:** Analyzing the prevention and control measures indicated that the strict social distancing implemented by the government effectively reduces the number of new cases and deaths. The curve of reproduction number also showed that effective reproduction number is still above one; hence, it is necessary to continue strict social distancing and control travelling to prevent causing another wave of outbreak especially in Persian New Year.

## 1. Background

COVID-19 is a global pandemic that emerged on December 31, 2019 in Wuhan, China [1] and has since then spread globally through the world. Iran reported the first confirmed cases of COVID-19 infections in Qom on February 19, 2020 [2]. The outbreak then quickly moved to all 31 provinces.

In response to the exponential increase in cases, the government closed all schools, universities and mosques. Friday prayers were cancelled and working hours were reduced by the government.

The closure of schools and universities, discontinuation of religious gatherings, prohibition of social events, travelling ban and the closure of government offices and many businesses in Nowruz holidays led to a decline of the effective reproduction number[3]. Consequently the number of new cases declined from the first peak in Nowruz. However, it increased once again and reached the highest value at the beginning of June and various interventions and policies implemented by government to control the spread of COVID-19. Nevertheless, another increase in the number of new cases occurs on October and the government implemented strict social distancing strategy and travelling ban to reduce the spread of the disease.

The aim of this study is to analyze the epidemic curves in 365 days coronavirus in Iran and to investigate the effect of social distancing and travelling on the spread of COVID-19 in Iran during one year spread.

## 2. Methods

We use the reported cases from February 20, 2020, to February 18, 2021, reported by MoHME [2]. We process the data and derive the features we need to analyze and then we drive the plots and analyze data.

### A) Correlation analysis

Correlation analysis is a statistical method applied to investigate the strength of the relationship between two or more variables. We use Pearson correlation coefficient to investigate the correlation between variables.

### B) Death rate

To calculate the death rate we use the number of daily deaths and daily new cases and calculate average daily death rate as the following:

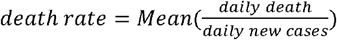

To investigate the effective reproduction number we use different methods as the following:

### C) JRC Method [4]

The basic reproduction number, *R*_O_, is the average number of secondary cases infected by a single infected person during infectious period in a complete susceptible population. *R*_O_ is defined [5] as the following:

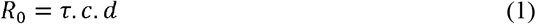

Where τ is the probability of infection per contact, c is the average contact rate, and d is the infectious period. *R*_O_ describes the average number of secondary infections when everyone is susceptible and there is no immunity in the population. However, in real world some people could be immune due to prior infection. Hence, the effective reproduction number, *R*_t_ , is defined as the average number of secondary cases per infectious case when there is some immunity in the population; and it can be estimated by the product of *R*_O_ and the fraction of the host population.

Considering the SIR (Susceptible-Infected-Removed) epidemic model [6], the change of the number of infected individuals is:

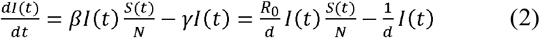

Where *s*(*t*) and *I*(*t*) are the number of susceptible and infected people respectively and *N* is the size of total population. *β* = *τ*.*c* is the infection rate, and 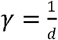 is the remove rate which is the inverse of infectious period.

In a complete susceptible population, 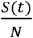 can be approximated by 1, then the solution of Eq.(1) is the following:

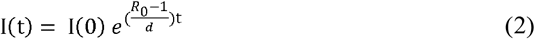

According to the definition of the effective reproduction number, *R*_t_, and Eq. (2) we have:

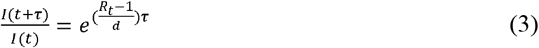

Hence, *R*_t_ can be calculated as the following:

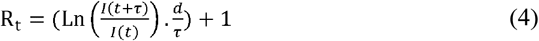

We used the number active infected individuals in each day I(t) and that of τ days after day t as *I* (*t*+ *τ*). We calculate R_t_ in each day of the outbreak considering d to be 6 days and τ to be 7.

### D) Robert Koch Institute method (RKI) method

In RKI [7] method (As cited in [4]), *R*_t_ is calculated as the total number of new cases during four consecutive days divided by that of four consecutive days prior to the days used in the denominator. We employ this method as the following:

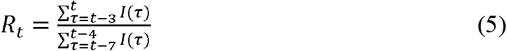

Where I(τ) is the number of new confirm cases in day τ of the outbreak.

## 3. Results

Fig. 1 shows the cumulative number of infected cases, recovered and deaths, and Fig. 2 shows the daily new confirmed cases.

**Fig 1.**
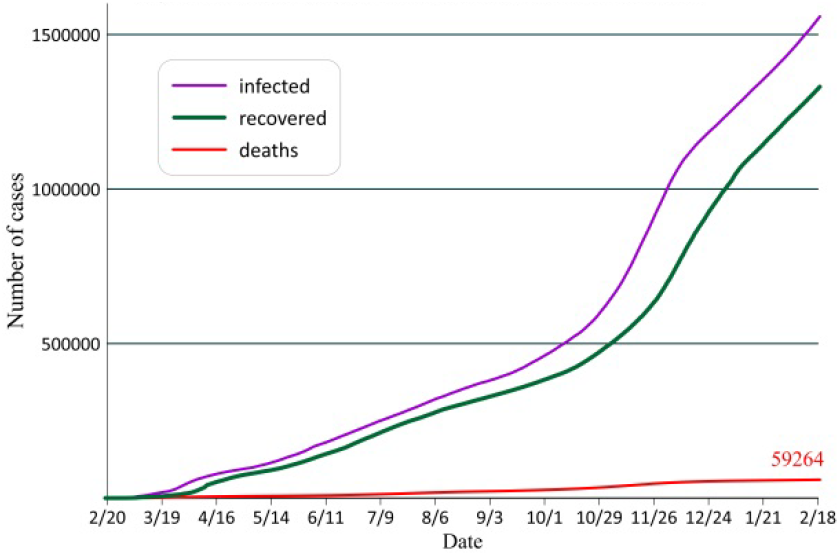
Cumulative number of infected, recovered and deaths

**Fig 2.**
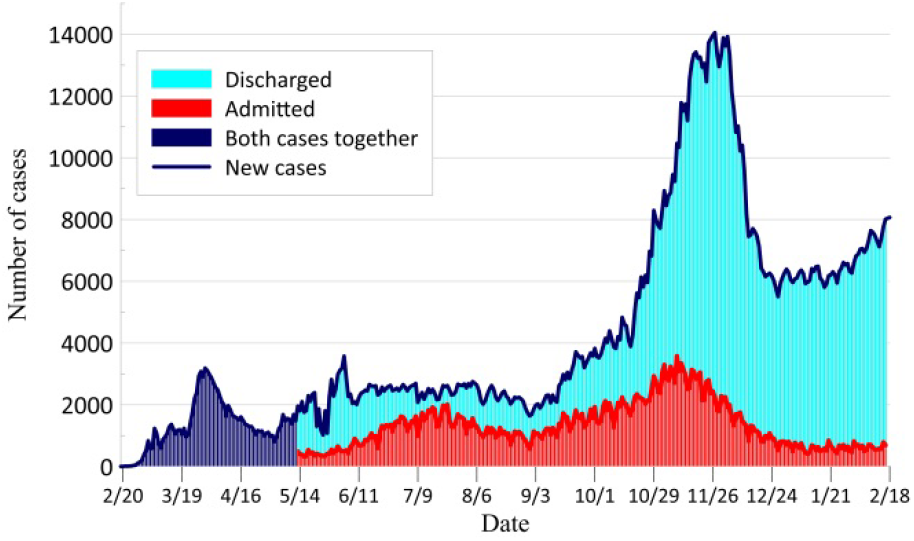
Daily new cases

Daily new cases are classified into two groups: admitted new cases which are hospitalized; and discharged new cases which are sent home. At the first two months of the outbreak, the summation of both cases was reported as daily new cases, however, from May 13, 2020 the number of discharged cases and admitted cases were reported separately. Hence, we show the number of new cases with three different colors in Fig. 2. The navy color shows the number of new cases until May 13, red and blue bars show the number of admitted and discharged new cases separately.

Fig. 3. shows the plot of daily new cases, daily recovered and daily deaths. We calculate the daily death rate using daily deaths divided by daily new cases. We use the same method to calculate the rate of daily positive tests per daily tests. Table 1. shows the statistics of daily death rate and daily confirmed rate. Fig. 4. shows the curve of daily death rate.

**Fig 3.**
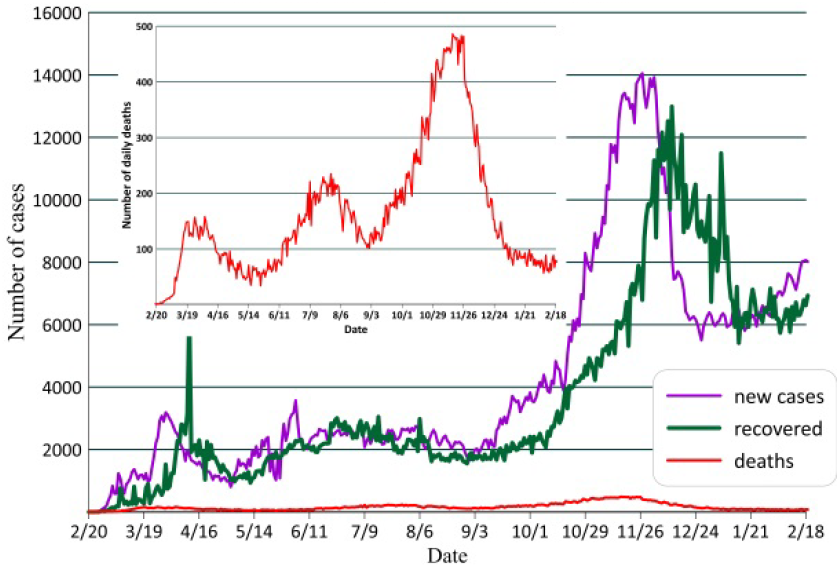
Daily new cases, recovered and deaths

**Table 1.** statistics of daily death rate and confirmed rate

|  |  |  |  |
| --- | --- | --- | --- |
| Count | 365 | Count | 317 |
| Mean | 0.049 | Mean | 0.139 |
| Min | 0.000 | Min | 0.056 |
| Max | 0.222 | Max | 0.354 |
| Variance | 0.001 | Variance | 0.004 |
| Standard Deviation | 0.028 | Standard Deviation | 0.065 |
| Standard Error of Mean | 0.001 | Standard Error of Mean | 0.004 |

A) death per new cases
B) confirmed cases per test

**Fig 4.**
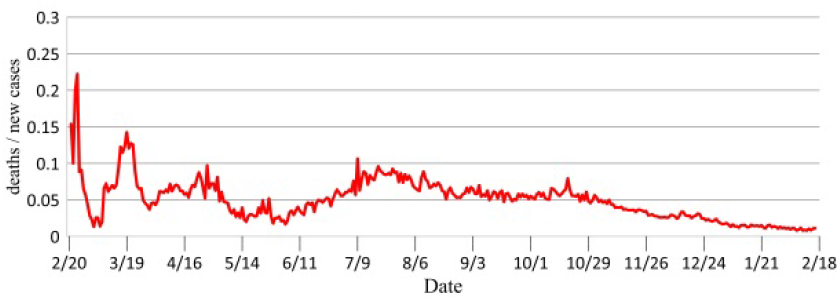
Daily deaths rate

The mortality rate of COVID-19 in Iran during one year outbreak can be calculated as cumulative deaths divided by cumulative number of infected cases which is 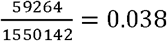. As can be seen in Fig. 4, the daily death rate is higher than its average and one year mortality rate in the early stages of outbreak and during March, April, July and August.

As can be seen in Table 1. The mean daily death rate of COVID-19 during 365 days is 4.9 percent. The statistical results also indicate that averagely 13.9 percent of daily tests results are positive.

Fig. 5 shows the curve of active infected cases in 365 days of outbreak. In Fig. 5. the size of plots shows the number of new cases and the colors indicate the strength of implementing social distancing. The red lines show the strong social distancing implementation, the blue lines show the medium social distancing policies and the navy lines indicates weak social distancing measures. School and university closure, travelling ban, Friday prayer cancelation, closure of government offices and closure of public places such as public libraries are considered as strong social distancing. Opening low risk business, opening government offices with reduced working hours, travelling allowance are considered as medium social distancing and opening all business and intervention as advice are considered as weak social distancing in Fig. 5. As can be seen in Fig. 5, strong social distancing such as travelling ban and government office closure reduces the number of new cases and active infected patients that consequently decrease daily deaths; however decreasing the limitations causes to increase the cases.

**Fig 5.**
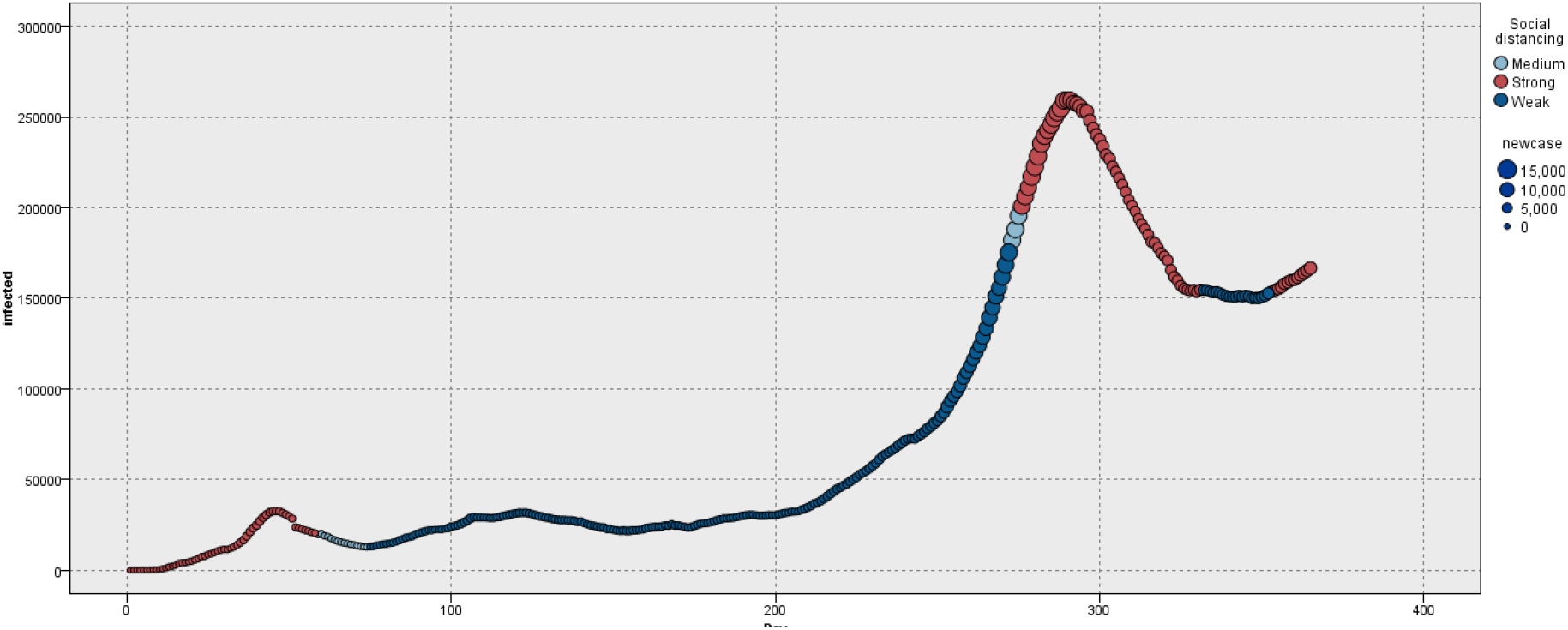
The curve of infected cases during 365 days of outbreak

Fig 6. shows the plot of effective reproduction number of COVID-19 during 365 days. As can been in Fig. 5 both JRC and RKI methods indicate that reproduction number were above four at the beginning of the outbreak and currently is above one. It is necessary to reduce and maintain the effective reproduction below one to control the disease.

**Fig 6.**
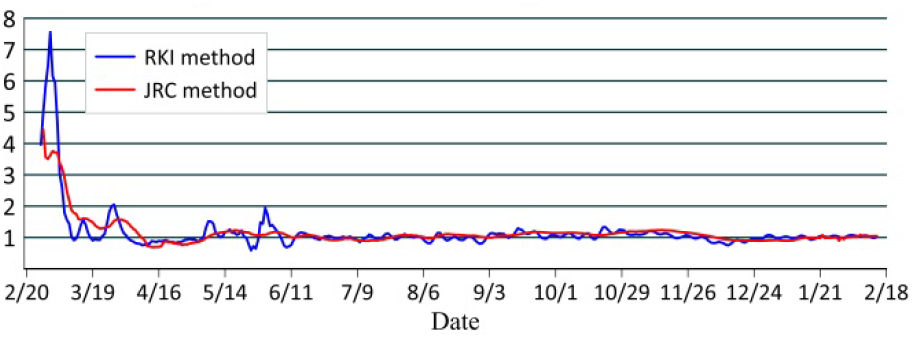
The curve of effective reproduction number

To analyze the correlation between variables we use Pearson correlations coefficient. Table 2. shows the correlation between variables.

**Table 2.**
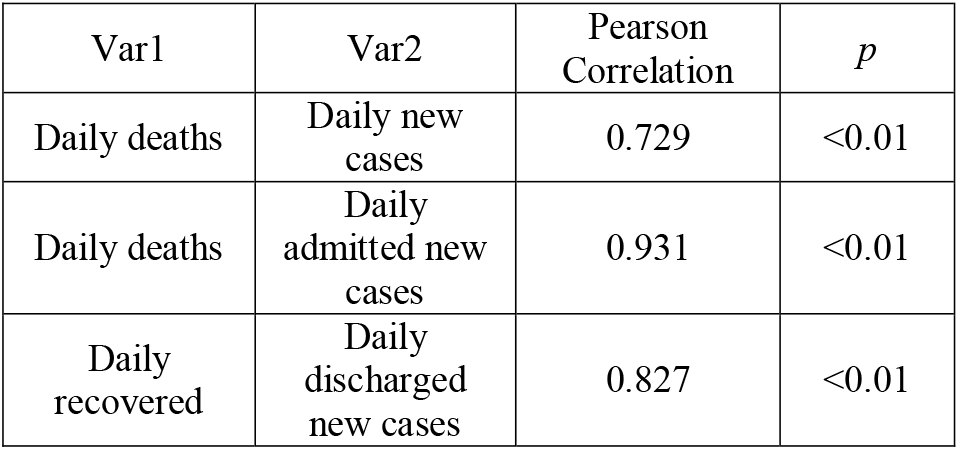
Correlation between variables

| Var1 | Var2 | Pearson Correlation | *p* |
| --- | --- | --- | --- |
| Daily deaths | Daily new cases | 0.729 | <0.01 |
| Daily deaths | Daily admitted new cases | 0.931 | <0.01 |
| Daily recovered | Daily discharged new cases | 0.827 | <0.01 |

As can be seen in table 2. There is a strong correlation between daily death and daily new cases specially admitted new cases.

## 4. Discussion

Various scholars have carried out research on COVID-19 in Iran [8][9][10]. Several researches have employed mathematical models to analyze the epidemic curve and forecast the epidemic trend of COVID-19 Iran [3][11][12]. Some studies have tried to investigate the relationship between COVID-19 and meteorological and climatological factors [13][14].

This study investigates the temporal characteristics of the COVID-19 outbreak in Iran. We used the official reported data to analyze the epidemic curves and to investigate the correlation between different epidemic parameters.

Fig. 2. and Fig. 3. illustrate the trend chart of daily confirmed cases in Iran. Since the epidemic outbreak, the first peak of daily confirmed cases was reached on March, 30, 2020 followed by continues decline until May, 2, 2020. Then, it increased again and reached the second peak, however, nonpharmaceutical interventions and the government advice/order controlled the disease.

At the beginning of September the number of daily confirmed cases started to increase once again. From mid-October the number of daily new cases rapidly increased and the government implemented strict social distancing measures such as office closure, travelling ban and traffic restriction.

As shown in Fig. 5. social distancing effectively reduced the number of active infected cases. Epidemic curves in Fig. 2. and Fig. 3. also indicate the same results. Importantly, our approach demonstrates that the number of daily deaths is associated with the number of new case. Hence, implementing control strategies to reduce the number of new cases can be an effective policy to reduce the deaths.

## 5. Conclusion

In this study we analyzed the epidemic curves and investigated the correlation between epidemic parameters of COVID-19 in Iran during 365 days of the outbreak. We also employed JRC and RKI methods to calculate the effective reproduction number of COVID-19 in Iran during one year outbreak.

The results of this study in analyzing the prevention and control measures implemented by government indicated that strong social distancing implemented by government has a great impact on the spread of the epidemic. Hence, it is necessary to continue social distancing and control travelling to prevent causing another wave of the outbreak.

## Data Availability

The data that support the findings of this study are available from the corresponding author upon reasonable request. The data were derived from the following public domain resource: https://behdasht.gov.ir/

## Abbreviations

Covid-19 coronavirus; effective reproduction number; data mining; Iran.

## Authors’ contributions

ES conducted analyses and wrote the manuscript; STA revised and edited the manuscript. All authors read and approved the final manuscript.

## Reference

[1] T. Chen, J. Rui, Q. Wang, Z. Zhao, J.-A. Cui, and L. Yin, “A mathematical model for simulating the phase-based transmissibility of a novel coronavirus,” Infect. Dis. Poverty, vol. 9, no. 24, p. 2020.01.19.911669, 2020.

[2] “Ministry of Health and Medical Education (MOHME).”.

[3] E. Sahafizadeh and S. Sartoli, “Epidemic curve and reproduction number of COVID-19 in Iran,” Journal of travel medicine, vol. 27, no. 5. 2020.

[4] A. Annunziato and T. Asikainen, “Effective Reproduction Number Estimation from Data Series,” 2020. [Online]. Available: https://publications.jrc.ec.europa.eu/repository/bitstream/JRC121343/r0_technical_note_v3.4.pdf.

[5] J. H. Jones, “Notes on R0,” Notes, pp. 1–19, 2007.

[6] W. O. Kermack and a. G. McKendrick, “A Contributions to the mathematical theory of epidemics,” Proc. R. Soc. London, vol. 115, no. 772, pp. 700–721, 1927.

[7] ROBERT KOCH INSTITUT, “Epidemiologisches Bulletin,” 2020. [Online]. Available: https://www.rki.de/DE/Content/Infekt/EpidBull/Archiv/2020/Ausgaben/17_20.pdf?blob=publicationFile.

[8] A. Raoofi, A. Takian, A. Akbari Sari, A. Olyaeemanesh, H. Haghighi, and M. Aarabi, “COVID-19 Pandemic and Comparative Health Policy Learning in Iran,” Arch. Iran. Med., vol. 23, no. 4, pp. 220–234, 2020.

[9] A. Zahiri, S. Rafieenasab, and E. Roohi, “Prediction of Peak and Termination of Novel Coronavirus Covid-19 Epidemic in Iran,” vol. 98, no. 51, pp. 1–13, 2020.

[10] M. Moghadami, M. Hassanzadeh, K. Wa, A. Hedayati, and M. Malekolkalami, “Modeling the corona virus outbreak in IRAN,” medRxiv, 2020.

[11] N. Ghaffarzadegan and H. Rahmandad, “Simulation-based estimation of the spread of COVID-19 in Iran,” medRxiv. pp. 1–19, 2020.

[12] A. Ahmadi, M. Shirani, and F. Rahmani, “Modeling and Forecasting Trend of COVID-19 Epidemic in Iran,” medRxiv, p. 2020.03.17.20037671, 2020.

[13] E. Sahafizadeh and S. Sartoli, “Rising summer temperatures do not reduce the reproduction number of COVID-19,” J. Travel Med., vol. 2020, pp. 1–3, Oct. 2020.

[14] M. Ahmadi, A. Sharifi, S. Dorosti, S. Jafarzadeh Ghoushchi, and N. Ghanbari, “Investigation of effective climatology parameters on COVID-19 outbreak in Iran,” Sci. Total Environ., vol. 729, 2020.

